# Psychosocial Factors of Stigma and Relationship to Healthcare Service among Adolescents Living With HIV/AIDS in Kano State, Nigeria

**DOI:** 10.1101/2020.12.22.20248702

**Authors:** Oladunni Abimbola Amos, Ayomide Busayo Sina-Odunsi, Boyiga Bodinga Nuga, Yusuff Adebayo Adebisi, Obasanjo Afolabi Bolarinwa, Adesina Adetoun Adeola, Don Eliseo Lucero-Prisno

**Affiliations:** Faculty of Pharmaceutical Sciences, Ahmadu Bello University, Zaria, Nigeria; Regional Office for the East and Horn of Africa, International Organization for Migration, United Nations Migration Agency, Nairobi, Kenya; International Organization for Migration, United Nations Migration Agency, Abuja, Nigeria; AB Global Health Initiative, Ogun state, Nigeria; Faculty of Pharmacy, University of Ibadan, Ibadan, Nigeria; Discipline of Public Health Medicine, College of Health Sciences, University of KwaZulu-Natal, South Africa; Department of Demography and Social Statistics, Faculty of Social Sciences, Obafemi Awolowo University, Nigeria; Global Health Focus, London, United Kingdom; Department of Global Health and Development, London School of Hygiene and Tropical Medicine, United Kingdom

**Keywords:** HIV/AIDS, Psychosocial factors, stigma, adolescents living with HIV, Nigeria

## Abstract

**Background:** Stigma associated with HIV shapes all aspect of prevention and treatment, yet there are limited data on how HIV-infected adolescents are affected by stigma. Stigma increases risk of psychological problems among HIV-infected individuals which can affect access to treatment and social support services. This study is aimed at identifying psychosocial factors of stigma and relationship to healthcare services among adolescents on antiretroviral therapy (ART) in Gwale Local Government Area (LGA) of Kano state, Nigeria.

**Methods:** A facility based cross-sectional survey was carried out from January 26 to February 28, 2020 across six health facilities providing ART service in Gwale local government. A structured interviewer-administered questionnaire was used to collect the data. ART clients attending clinics were interviewed following an informed verbal consent. Descriptive statistics was used to summarize the data and results are presented using simple frequency tables and percentages. Upon completion of univariate analysis, the data was analyzed at the bivariate level using chi-square test to determine associations between different variables.

**Results:** One hundred and eight (108) clients voluntarily participated in the study of which 54 (50%) are male respondents and 54 (50%) are female respondents. Under the internalized stigma item, 67% of HIV-infected adolescents who have lost father or mother to AIDS reported feeling less valuable than other children who are not infected with HIV. Under the perceived stigma items, 86% of participants who have lost father or mother to AIDS reported to have excluded themselves from health services and social activities in the last twelve months due to fear of being insulted. Under the experienced stigma items, 62% of participants who have lost father or mother to AIDS reported to have been avoided by friends and colleagues in the last twelve months.

**Conclusion:** The study revealed that loss of intimate relation (father or mother) to AIDS and equal treatment with other HIV negative siblings were found to be significantly associated with the three forms of stigma (internalized stigma, perceived stigma and experienced stigma) including access to healthcare services. There is need for social and psychological support programs among HIV-infected adolescents.

## INTRODUCTION

The emergence of Human Immunodeficiency Virus (HIV) has brought about significant health challenges in children and adolescents across the globe (UNICEF 2018) manifesting as biological, psychological, and social problems. While the introduction of antiretroviral therapy (ART) has offered hope to children living with HIV (Patel et al. 2005; Collins et al. 2010), stress associated with HIV diagnosis is a challenge that needs to be overcome. Globally, an estimated 38 million people were living with HIV in 2019, out of which 2.8 million were children age 0-19 years (UNICEF 2020). In 2019, UNAIDS and the National Agency for the Control of AIDS estimate that there are 1.9 million people living with HIV in Nigeria (UNAIDS 2019). Although advances have been made on early childhood survival among HIV-infected children, progress has been slow among adolescent population; between 2003 and 2019, the number of AIDS-related deaths annually among children has declined by 74.0% while the number of AIDS-related deaths annually among adolescents (10-19 years) have only decreased by 6.0% (UNICEF 2020). It was estimated, in 2014, that 50.0% of orphans with HIV have grown into adolescence (Monasch and Boerma 2004) and 740,000 adolescents could become additionally infected between 2016 and 2030 (UNICEF 2016).

The psychosocial wellbeing of children is crucial to achieving and maintaining optimum therapy and quality of life (Anouk et al. 2013). This has been significantly impacted due to the challenges of having to deal with a disease that is associated with increase in mental health problems (Scharko 2006). These challenges include mental distress due to loss of caregivers or family members to AIDS, internalized stigma, discrimination, disclosure issues and difficulties associated with antiretroviral therapy (ART) adherence (McCleary-Sills et al. 2013; Petersen et al. 2010). Evidence has revealed negative correlation between HIV infection and cognitive development and function in children, highlighting the critical need to address this challenge during early stage of childhood development (Sherr 2011; Nichols et al. 2013). HIV positive children are at risk of developing behavioral problems as they grow into adolescent stage (Mellins et al. 2008) which subsequently impact adherence to antiretroviral therapy (ART), sexual behavior, treatment outcome and overall wellbeing (Lowenthal at al. 2012).

Improved health outcome and quality of life in HIV infected children has been associated with psychosocial wellbeing (Mellins et al. 2004). Hence, a need to identify factors that contribute to resilience in HIV-infected orphans and vulnerable children has been promoted among scholars and practitioners (Skordal, 2012). Study has identified age, cognitive ability, sense of purpose and belief in positive future as factors that contribute to resilience (Goldstein and Brooks 2013). Factors at individual level, family level (parents or caregivers wellbeing), institutional level (access to education, health services) and societal level (stigma and discrimination) forms a complex interplay that shapes resilience in HIV-infected children (McCleary-Sills et al. 2013). Study conducted on psychosocial wellbeing of children who have loss parent or caregiver to HIV or affected by HIV has examined factors that impacts their unique vulnerabilities (Richter et al. 2006). These vulnerabilities are often interplayed between managing illness and loss of parent which is expanded by associated stigma. Children and adolescents living with HIV are commonly faced with the challenge of having to deal with specific psychosocial issues in addition to coping with the disease. Therefore, addressing these psychosocial needs should not only focus on factors associated with chronic diseases but should also consider common HIV-related factors such as stigma and discrimination, disclosure challenges, challenges associated with treatment access and retention in care. This paper aimed to explore the psychosocial factors associated with stigma and how they affect access to HIV health service delivery among adolescents living with HIV/AIDS in Gwale Local Government Area (LGA) of Kano state, Nigeria.

## METHODS

### Study design and setting

This study was a health facility-based cross-sectional survey of adolescents living with HIV/AIDS (ALWHA) in Gwale Local Government Area of Kano state from January 26 to February 28, 2020. Eligible participants were ALWHA enrolled on antiretroviral therapy (ART) who gave voluntary informed consent to participate in the study. Only participants (age 10 to 20 years) who attended ART clinics were recruited successively and interviewed by health workers during their visit to the clinics in the selected health facilities.

### Study Area

A health facility based cross-sectional survey was carried out in six health facilities providing ART services for People Living with HIV/AIDS (PLWHA) across Gwale Local Government Area (LGA). Gwale LGA is located within Greater Kano City with an area of 18km^2^ and population of 362,059 according to the 2016 census. There are twelve primary healthcare facilities distributed across twelve wards in the area. However only six healthcare facilities (Gwale Primary Health Center (PHC), Dorayi Karama PHC, Jaen PHC, Kabuga PHC, Unguwar Dabai PHC, and Pilot Kwanar Ganduje) were providing ART services.

### Sample Size Determination

Number of ALWHA who were enrolled on ART was obtained from the Department of Health at Gwale Local Government Area Secretariat. Based on the estimated population of 148 ALWHA attending ART clinics and using assumption of 95.0% confidence level and 5.0% margin of error, a sample size of 108 was obtained using Yamane sample size formula (Yamane, 1967).

### Sampling and data Collection Procedure

The questionnaire was distributed and administered through trained interviewers who are health workers across the six health facilities that provide ART. Health workers who interviewed the study participants were recruited across the six health facilities. Interviewers who understood the local language were recruited for the study in order to properly interpret the questionnaire in instances where participants do not understand English. ALWHA who are eligible for the study were recruited by trained health workers at the selected health facilities in the local government and consecutive sampling technique was used to enroll participants. The questionnaire was administered after voluntary verbal informed consent was obtained to signify intention to participate in the study. Anonymity and confidentiality of response were assured, while participation was entirely voluntary. The principal investigator supervised the data collection process on daily basis. No incentive was given to our respondents for participating in the study.

### Data Collection Instrument, Pretest and Content Validation

A structured questionnaire was adapted from HIV stigma index evaluation survey conducted in six cities in Iran (SeyedAlinaghi et al. 2013). The questionnaire was pretested among five ALWHA during clinic visit at Maternal Child Health Center in Gwale local government and these respondents were not included in the main study. The feedback obtained from the pretest led to minor modification of the questionnaire. The questionnaire was assessed for ambiguity and content validity by two researchers in order to ascertain the comprehensiveness of the question items. The questionnaire was grouped into four sections. Section A captured sociodemographic characteristics and psychosocial factors, Section B comprised questions on internalized stigma, Section C was on experienced stigma and Section D comprised questions on perceived stigma.

### Data Analysis

Data was checked for appropriateness of responses and completeness before it was entered into Microsoft office 2013 spreadsheet. After completing the data entry, the data were transferred into SPSS version 20 for coding, checking, and cleaning then transferred to STATA version 13 for analysis. Descriptive statistics were used to summarize the data and the results were presented using simple frequency tables and percentages to see the overall distribution of the study subject with the variable under study was done. Upon completion of univariate analysis, the data was also analyzed at the bivariate level of analysis using Chi-Square test. This statistic was used to test the associations between different variables. The criteria for statistical significance was set at p-value less than 0.05.

### Ethical Consideration

Ethical approval for the study was obtained from Kano State Ministry of Health Institutional Review Board. All study participants were informed about the purpose of the study and voluntary verbal consent was obtained from all participants.

## RESULTS

A total of 108 study participants (50.0% men and 50.0% women) responded to the questionnaire survey. **Table 1** outlines the socio-demographic characteristic of the respondents; 16.7% of the respondents were within the age of 10-14 while most of the respondents within the age 15-19 were 78.7% and only 4.6% were above 20 years of age. Those who belong to the Hausa and Fulani ethnic group were 53.7% and 38.9% respectively, while 7.4% represents those from other ethnic group.

**Table 1:** Percentage Distribution of Participants by Socio-Demographic Characteristics.

| **Variable (n=108)** | **Frequency (%)** |
| --- | --- |
| **Sex** |  |
| Male | 54 (50.0) |
| Female | 54 (50.0) |
| **Age** |  |
| 10-14 | 18 (16.7) |
| 15-19 | 85 (78.7) |
| ≥20 | 5 (4.6) |
| **Ethnicity** |  |
| Fulani | 42 (38.9) |
| Hausa | 58 (53.7) |
| Others | 8 (7.4) |
| **Marital Status** |  |
| Single | 67 (62.0) |
| Married | 37 (34.3) |
| Separated | 4 (3.7) |
| **Educational Attainment** |  |
| No Formal Education | 31 (28.7) |
| Primary Education | 20 (18.5) |
| Secondary Education | 32 (29.6) |
| Postsecondary Education | 25 (23.2) |

**Table 2** shows the percentage distribution of participants by psychosocial factors. The results show that less than half (47.3%) of the respondents have parental relationship with caregiver, 19.4% have spousal relationship with caregiver and 33.3% of the respondents have some other kind of relationship with the caregiver. As regards to respondents’ current state of mind under the internalized stigma items, 61.1% percent of them were most times anxious/afraid, 34.3% were most times happy/stable and only 4.6% were depressed/feel like committing suicide. Among respondents who reported perception of being stigmatized under the perceived stigma items, 34.3% reported not to be anxious/afraid, 29.6% reported fear of death because disease has no cure, 23.1% reported fear of being segregated, 10.2% report uncertainty about marriage/future intimate relationship and only 2.8% reported to be fed up. With respect to experienced stigma, result shows that 65.7% of the respondents have been avoided by friends/colleagues most of the time during social interaction in the past 12 months and 34.3% of the respondents reported otherwise.

**Table 2:** Percentage Distribution of Participants by Psychosocial Factors.

| **Variables (n=108)** | **Frequency (%)** |
| --- | --- |
| **Caregiver' relationship** |  |
| Parent | 51 (47.3) |
| Spouse | 21 (19.4) |
| Others | 36 (33.3) |
| **HIV disclosure to friends/neighbour** |  |
| No | 24 (22.2) |
| Yes | 84 (77.8) |
| **Loss of intimate relation (father or mother) to AIDS** |  |
| No | 49 (45.4) |
| Yes | 59 (54.6) |
| **Received equal treatment with HIV negative siblings in last 12months** |  |
| No | 36 (33.3) |
| Yes | 72 (66.7) |
| **Allowed to share utensils with HIV negative siblings** |  |
| No | 41 (38.0) |
| Yes | 67 (62.0) |
| **Internalised Stigma** |  |
|  | **Frequency (%)** |
| **Feeling about your current state of mind** |  |
| Anxious/afraid most times | 66 (61.1) |
| Depressed/feel like committing suicide | 5 (4.6) |
| Happy/stable most times | 37 (34.3) |
| **Feeling about your general health condition** |  |
| Fair | 19 (17.6) |
| Good | 86 (79.6) |
| Poor | 3 (2.8) |
| **Less Valuable than those not infected** |  |
| No | 65 (60.2) |
| Yes | 43 (39.8) |
| **Denied/excluded yourself from access to health services in the last 12months** |  |
| No | 31 (28.7) |
| Yes | 77 (71.3) |

| Perceived Stigma |  |
| --- | --- |
|  | Frequency (%) |
| **What is your reason for being anxious/afraid?** |  |
| Fear of being segregated | 25 (23.1) |
| Fear of death because disease has no cure | 32 (29.6) |
| Fed up | 3 (2.8) |
| Not anxious/afraid | 37 (34.3) |
| Uncertainty about marriage/future intimate relationship | 11 (10.2) |
| **Reason for denying/excluding yourself** |  |
| Fear of being gossiped about | 34 (31.5) |
| Fear of being insulted | 22 (20.4) |
| Fear of being rejected/harassed | 21 (19.4) |
| Never denied myself | 31 (28.7) |
| Experienced Stigma |  |
|  | Frequency (%) |
| **Been avoided by friends/colleagues** |  |
| No | 37 (34.3) |
| Yes | 71 (65.7) |
| **Has anyone ever referred to you as irresponsible/guilty?** |  |
| No | 22 (20.4) |
| Yes | 86 (79.6) |
| **Has anyone ever referred to your relative as being irresponsible?** |  |
| No | 38 (35.2) |
| Yes | 70 (64.8) |
| **Have you ever been denied service in a health organization in the last 12 months?** |  |
| No | 51 (47.2) |
| Yes | 57 (52.8) |

**Table 3** shows the association between psychosocial and internalized stigma of the study respondents. Over three-fifths (67.4%) of the respondents who have lost intimate relation (father or mother) to AIDS felt that they are less valuable to those who are not infected with HIV. Among those who received unequal treatments with their siblings, about 9 in 10 persons have also felt been less valuable than those who are not infected with HIV. Both psychosocial variables used in this study shows association with internalized stigma with *p-value* less than *0*.*05*.

**Table 3:** Association between psychosocial and internalized stigma.

| Lost father or mother to AIDS | Less Valuable than those not infected |  |
| --- | --- | --- |
|  | No | Yes |
| No | 53.8% | 32.6% |
| Yes | 46.2% | 67.4% |
| $X^2 = 4.73$ $p\text{-value}=0.03^*$ | | |
| Equal treatment with HIV-siblings |  |  |
| No | 50.8% | 7.0% |
| Yes | 49.2% | 93.0% |
| $X^2=22.33$ $p\text{-value}=0.00^*$ | | |

**Table 4:** psychosocial characteristics of the respondents and perceived stigma shows that among those who have lost father or mother to AIDS, more than four-fifths (86.4%) excluded themselves from health services and social activities in the last twelve months because they felt they will be insulted. Also, among those who had experienced equal treatment with siblings almost 9 in every 10 persons excluded themselves from health services and social activities in the last twelve months because of the fear of gossip as shown in table 4 below. Both psychosocial variables used in this study shows association with respondents’ perceived stigma with *p-value* less than *0*.*05*.

**Table 4:** Association between psychosocial and perceived stigma.

| Lost father or mother to AIDS | Reason for excluding yourself from health services and social activities |  |  |  |
| --- | --- | --- | --- | --- |
|  | Gossip | Insult | Reject | Never |
| No | 38.2% | 13.6% | 38.1% | 80.6% |
| Yes | 61.8% | 86.4% | 61.9% | 19.4% |
| $X^2=25.65$ $p\text{-value}=0.00^*$ | | | | |
| Equal treatment with HIV-siblings |  |  |  |  |
| No | 11.8% | 22.7% | 52.4% | 51.6% |
| Yes | 88.2% | 77.3% | 47.6% | 48.4% |
| $X^2= 16.32$ $p\text{-value}=0.00^*$ | | | | |

**Table 5** shows the association between psychosocial and experienced stigma of the study respondents. Slightly above three-fifths (62.0%) of respondents who have experienced loss of intimate relation (father or mother) to AIDS reported to have been avoided by friends or colleagues. Among those who experienced equal treatment with siblings from their parents, more than four-fifths (84.5%) respondents have reported to have been avoided by friends or colleagues. Both psychosocial variables used in this study shows association with respondents experienced stigma with *p-value* less than *0*.*05*.

**Table 5:** Association between psychosocial and experienced stigma.

| Lost intimate to AIDS | Avoided by friends or colleagues |  |
| --- | --- | --- |
|  | No | Yes |
| No | 59.5% | 38.0% |
| Yes | 40.5% | 62.0% |
| $X^2 = 4.51$ $p\text{-value}=0.03^*$ | | |
| Equal treatment with HIV-siblings |  |  |
| No | 67.6% | 15.5% |
| Yes | 32.4% | 84.5% |
| $X^2=29.68$ $p\text{-value}=0.00^*$ | | |

Socio-demographic characteristics of respondents and healthcare services access is shown in **Table 6**. All the selected socio-demographic variables showed an association with being denied access to healthcare service with *p-value* less than *0*.*05* except respondents’ ethnic groups.

**Table 6:** Socio-demographic characteristics associated with HIV care services access.

|  |  |  |
| --- | --- | --- |
| Gender | Denied of access to healthcare service in the last 12months |  |
|  | No | Yes |
| Female | 13.7% | 82.5% |
| Male | 86.3% | 17.5% |
| $X^2 = 50.86$ $p\text{-value}=0.00^*$ | | |
| Age |  |  |
| 10-14 | 13.7% | 19.3% |
| 15-19 | 78.5% | 79.0% |
| 20 years & above | 7.8% | 1.7% |
| $X^2=22.33$ $p\text{-value}=0.00^*$ | | |
| Ethnicity |  |  |
| Fulani | 31.4% | 45.6% |
| Hausa | 60.8% | 47.4% |
| Others | 7.8% | 7.0% |
| $X^2=2.33$ $p\text{-value}=0.31$ | | |
| Marital status |  |  |
| Married | 5.9% | 59.7% |
| Separated | 4.0% | 3.5% |
| Single | 90.1% | 36.8 % |
| $X^2=35.08$ $p\text{-value}=0.00^*$ | | |
| Literacy level |  |  |
| No formal education | 7.8% | 47.4% |
| Post-secondary | 39.2% | 8.7% |
| Primary education | 11.8% | 24.6% |
| Secondary education | 41.2% | 19.3% |
| $X^2=32.15$ $p\text{-value}=0.00^*$ | | |
| Family type |  |  |
| Monogamous | 43.1% | 63.2% |
| Polygamous | 56.9% | 36.8% |
| $X^2=4.33$ $p\text{-value}=0.03^*$ | | |

**Table 7** shows relationship between psychosocial characteristics of respondents and whether they have been denied access to healthcare services in the last twelve months; 70.2% of respondents who have lost an intimate relation to AIDS before reported to have been denied an access to healthcare services in the last twelve months while 9 in every 10 of those who experienced equal treatment with their siblings reported to have been denied access to healthcare services in the last twelve months. Both psychosocial characteristics of the respondents were associated with access to HIV care services with *p-value* less than *0*.*05*.

**Table 7:** Psychosocial characteristics associated with Access to HIV care services.

| Lost intimate relation to AIDS | Denied of access to HIV care services |  |
| --- | --- | --- |
|  | No | Yes |
| No | 62.8% | 29.8% |
| Yes | 37.2% | 70.2% |
| $X^2 = 11.77$ $p\text{-value} = 0.00^*$ | | |
| Equal treatment with HIV-siblings | Denied of access to health facilities |  |
|  | No | Yes |
| No | 62.7% | 7.0% |
| Yes | 37.3% | 93.0% |
| $X^2 = 37.61$ $p\text{-value} = 0.00^*$ | | |
| Lost intimate relation to AIDS | Denied self of access to HIV care services |  |
|  | No | Yes |
| No | 80.7% | 31.2% |
| Yes | 19.3% | 68.8% |
| $X^2 = 21.8285$ $p\text{-value} = 0.00^*$ | | |
| Equal treatment with HIV-siblings | Denied self of access to HIV care services |  |
|  | No | Yes |
| No | 51.6% | 26.0% |
| Yes | 48.4% | 74.0% |
| $X^2 = 6.5379$ $p\text{-value} = 0.01^*$ | | |

## DISCUSSION

Children and adolescents living with HIV and AIDS can conceive negative beliefs about HIV and stigmatize themselves which is referred to as internalized or self-stigma (Nikus et al. 2016). Fear of being stigmatized in school, neighborhood and other social gathering may arise which is referred to as perceived stigma. HIV stigma can be worsened when children and adolescents are ostracized or exposed to acts of discrimination and abuse. This is termed enacted or experienced stigma. Hence, this health facility-based cross-sectional study was conducted to determine the prevalence of the three forms of stigmas and to identify related psychosocial factors and how these factors affects access to healthcare services among HIV-infected adolescents.

According to this study, anxiety and fear was found to be significantly high; 61.1% of participants reported feeling anxious and afraid most of the time under the internalized stigma items and fear of death because the disease has no cure (29.6%), fear of being segregated (23.1%) and uncertainty about marriage (10.2%) were identified reasons for being anxious and afraid. This may be due to negative beliefs about their state of health and incertitude associated with being infected with HIV. There is need to improve coping strategies through emotional self-regulation in HIV-infected adolescents. Study have shown that children with better emotional regulation are more positive and are more likely to utilize available resources (Cooke et al. 2019). Findings from this study also revealed that 71.3% of adolescents have denied or excluded themselves from healthcare services in the last twelve months periods and the reasons for this occurrence were identified to be fear of being gossiped about (31.5%), fear of being insulted (20.4%) and fear of being rejected or harassed (19.4%). This is consistent with a review and study conducted which revealed that internalized stigma undermines treatment adherence and retention in care (Katz et al. 2013; Valverde et al. 2018). This situation can result in poor mental health, less social support, and more HIV-related symptoms in HIV-infected adolescents.

Stigma-related experiences including discrimination, physical violence and social rejection promotes psychosocial consequences in PLWHA which can be detrimental to their treatment behavior (Clark et al. 2003; Major and O’Brien 2005). Participants in this study reported experiences of being stigmatized and discriminated against in public. These experiences included being avoided by friends/colleagues (65.7%) and being denied service in an organization (52.8%). Verbal insults are also reported among participants including awareness of being referred to as irresponsible/guilty due to HIV seropositive status (79.6%) and awareness of relatives being referred to as irresponsible due to HIV-positive status (64.8%). Similar study conducted in Ethiopia showed that more than half of respondents (58.5%) indicated awareness of being gossiped about, while 39% of respondents reported being verbally insulted or harassed due to their HIV-status (Nikus et al. 2016). Higher prevalence of discrimination and abuse may be due to dynamics of sociocultural context such as linking PLWHA with immoral practices and poor level of awareness about stigma and potential harmful effect on individuals, families, community, and the healthcare system.

Our result showed that loss of father or mother to AIDS and equal treatment with other HIV-negative siblings were significantly related to internalize stigma, perceived stigma and experience stigma. In relation to internalized stigma, three out of five respondents who have lost father or mother to AIDS felt less valuable than other children who are not infected with HIV. This finding resonates with Dowdney, 2000 which highlighted that parental bereavement irrespective of its cause can put children at risk of internalizing problems such as depression, anxiety, withdrawal, and low self-esteem. The loss of one or both parents generally worsens psychological symptoms in children which can be deleterious to their wellbeing (Atwine et al. 2005; Cluver et al. 2007; Li et al. 2009). Among participants who did not receive equal treatment with other HIV negative siblings, nine in ten of them felt less valuable than other children who are not infected. Studies have suggested that HIV and AIDS stigma experienced at caregiver level (irrespective of their serostatus) negatively affects HIV-infected children (Rebhun 2004; Mawn 1999; Poindexter 2002). This indicates that caregivers’ discriminatory attitude towards HIV-infected children and adolescents can trigger psychosocially constructed view about HIV which can further be assimilated and internalized. This can have adverse effect on their mental health and health seeking behavior as they develop into adulthood.

In relation to perceived stigma, more than half of respondents who have lost father or mother to AIDS have excluded themselves from health services and social activities in the last twelve months due to perception of being insulted or being rejected or being gossiped about. This finding is concordant with a study conducted in Cambodia which showed that out of the 146 (79.8%) children on ART who have lost one or both parents to AIDS, a total of 88 (60.3%) felt they are being negatively treated including being rejected by others, not invited to social activities and exclusion from games (Barennes et al. 2014). This may be due to anxiety experienced during parent deteriorating health condition and trauma following the death of a parent (Pivnic and Villegas, 2000). AIDS-orphaned children may not only be traumatized by the bereavement of parents but also through absence of appropriate parental guidance through important life stages of socialization and identity formation amidst AIDS associated stigma and discrimination (Paul, 2009). There is need for more studies to evaluate broader risk of perceived stigma in HIV-infected adolescents. On the other hand, more than half of respondents have excluded themselves from health services and social activities in the last twelve months among those who received equal treatment with other HIV negative siblings in the last twelve months due to perception of being gossiped about or insulted. These perceptions of being stigmatized in public may have arose from other levels of influences that are beyond family level including individual, institutional, and societal level.

In relation to experienced stigma, more than half (62.0%) of respondents who have lost mother or father to AIDS have reported being avoided by friends and colleagues in the last twelve months compared to non-orphaned adolescents (38.0%). Consistent with a study conducted in Cape Town, South Africa which showed that adolescents who are orphaned by AIDS reported higher level of stigma than non-orphaned adolescents (Cluver et al. 2008). This situation can lead to poorer psychological outcomes including anxiety, depression, and post-traumatic stress, reduced level of social support and increase sense of isolation (Cluver et al. 2008).

In this study, sex, family size and age of respondents were significantly associated with being denied access to health care. Female adolescents have significantly experienced being denied access to health care in the last twelve months. This finding resonates with a study conducted in Jimma town, Southwest Ethiopia which revealed that although being denied access to dental/health services was scored lowest among the experienced stigma items, HIV-infected females respondents had a significantly higher mean score of experienced stigma compared to the male respondents (Nkus et al. 2016). The reason for this can be attributed to the negative constructed view about females living with HIV/AIDS. Women tend to suffer more from HIV-related stigmas because the community views them as being promiscuous in the past when they are infected with HIV. Monogamous family type was also significantly associated with being denied access to health care.

AIDS-orphaned children are likely to experience hostility from extended families and the community including being rejected or denied access to healthcare (Bauman et al. 2002; Ntozi & Mukiza-Gapere, 1995). Our finding shows that out of adolescents who have lost father or mother to AIDS, 70.2% have reported been denied access to health care in the last twelve months. This may be due to irrational fear of being infected with HIV among healthcare workers. This situation can exacerbate the psychological distress imposed by death of parent thereby having negative consequence on their retention in care and adherence to ART. Consistent with our finding in this study which revealed that 68.8% of adolescents who have lost father or mother to AIDS reported to have withdrawn themselves from HIV care services in the last twelve months.

## STRENGTH AND LIMITATION OF THE STUDY

The study had high completion rate and provides an understanding of stigma characteristics and retention in care among ALWHA, thereby bringing to focus the contemporary areas of further study and improvement in practice. The high response rate was achieved because questionnaire interview was conducted during participants’ clinic visits. However, this study is not without its limitation. One of the limitations was the use of closed-ended (yes/no) questions; a more accurate representation of questions would be the use of stigma scale. More so, the study was conducted in one local government area perhaps an expanded knowledge of stigma characteristics might be obtained if the study was conducted in more ART clinics in other local governments.

## CONCLUSION

This study shows that verbal and social stigma, discrimination in AIDS orphaned and non-orphaned adolescents living with HIV/AIDS remains a significant issue in Kano state, Nigeria. These unhealthy experiences have negative impact on treatment adherence. Social support for HIV-infected adolescents helps them cope with stigma and discrimination. There is need for psychosocial support programmes for AIDS-orphaned and non-orphaned adolescents on ART. Context specific stigma reduction strategies and psychological support should target multiple levels of influence including intrapersonal, interpersonal and structural level factors in order to build resilience in HIV-infected adolescents. Education programs should target prevention of HIV specific stigma through the media, schools, and hospitals in Kano state. More studies are required to characterize stigma and to explore interventions that promotes patient-healthcare provider relationship and retention in care among HIV-infected adolescents in Kano State.

## Data Availability

Data will be made available on request

